# Validity of gait parameters of healthy young adults using a motion-sensor-based gait analysis system (ORPHE ANALYTICS) during walking and running

**DOI:** 10.1101/2022.10.17.22281166

**Authors:** Yuki Uno, Issei Ogasawara, Shoji Konda, Natsuki Yoshida, Akira Tsujii, Ken Nakata

## Abstract

**Background:** Motion sensors are widely used for gait analysis. ORPHE ANALYTICS is a motion-sensor-based gait analysis system. The validity of commercial gait analysis systems is of great interest to clinicians because calculating position/angle-level gait parameters using motion sensor data potentially produces an error in the integration process; moreover, the validity of ORPHE ANALYTICS has not yet been examined.

**Research question:** How valid are the position/angle-level gait parameters calculated using ORPHE ANALYTICS relative to those calculated using conventional optical motion capture?

**Methods:** Nine young adults performed gait tasks on a treadmill at speeds of 2–12 km/h. The motion sensors were mounted on the shoe midsole (plantar-embedded) and shoe instep (instep-mounted). The three-dimensional marker position data of the foot as well as the acceleration and angular velocity data of the motion sensors were collected. The position/angle-level gait parameters were calculated from motion sensor data obtained using ORPHE ANALYTICS and optical motion capture data. Intraclass correlation coefficients [ICC_(2,1)_] were calculated for relative validities, and Bland–Altman plots were plotted.

**Results:** Eight items, namely, stride duration, stride length, stride frequency, stride speed (plantar-embedded), vertical height (plantar-embedded), stance phase duration, swing phase duration, and sagittal angle_IC_, exhibited excellent relative validities [ICC_(2,1)_ > 0.9]. In contrast, the sagittal angle_TO_ demonstrated good relative validity [ICC_(2,1)_ = 0.892–0.833], while the frontal angle_IC_ exhibited moderate relative validity [ICC_(2,1)_ = 0.566–0.627].

**Significance:** ORPHE ANALYTICS, a motion-sensor-based gait analysis system, was found to exhibit excellent relative validity for most gait parameters. This finding suggests its feasibility for gait analysis outside the laboratory setting.

**Highlights:**

- Gait-parameter validities were examined for treadmill-based gait tasks at 2–12 km/h.
- Most gait parameters showed excellent relative validity with optical motion capture.
- Shoe midsole-embedded sensors had higher validities than instep-mounted sensors.
- ORPHE ANALYTICS is potentially useful in clinical measurements.

## Introduction

Motion sensors (inertia sensors) have become widely used for the motion measurement of any type of object owing to their rapid technological development and have recently been applied to gait analysis[1]. Motion sensors are highly compact and lightweight, allowing their attachment to the body without spatially restricting the object of measurement. These features potentially overcome the significant limitations of optical cameras and reflective-marker-based motion capture systems commonly used in motion measurement, which are only feasible in a limited laboratory space[2]. The motion sensor will expand the range of applications, such as a more natural gait assessment in daily life and frequent measurements without visiting a specific facility[3].

Several commercial packages that take advantage of motion-sensor compactness and lightweight are currently available[4–7]. One commercial product is ORPHE ANALYTICS (ORPHE Inc., Tokyo, Japan), which is a gait analysis system that can evaluate position/angle-level gait parameters (such as stride length and foot-ground angle) using the acceleration and angular velocity data of shoes during walking and running. These data are collected using a 20-g sensor mounted on a shoe’s midsole or instep. Therefore, provided individuals wear their smart shoes, assessing their gait patterns without constraining the measurement environment is possible.

Position/angle-level gait parameters are commonly utilised in conventional gait assessments[8] because conventional gait analyses have frequently used optical motion capture, which can measure the position and angle of the targeted body segment or shoe. Furthermore, position/angle-level gait parameters are easier to understand than derivative quantities, such as accelerations and angular velocities. This background probably motivates the calculation of position/angle-level gait parameters in motion-sensor-based gait analysis. However, because motion sensors cannot directly measure the position/angle-level kinematic properties, numerical integration of the measured acceleration or angular velocity data is necessary. The numerical integration process poses certain challenges, such as the production of numerical errors. These errors are affected by sensor specifications[9,10] and mounting position[11,12]. Therefore, the use of motion-sensor-based commercial gait analysis systems to determine how consistently position/angle-level gait parameters are obtained from motion sensors is of great interest. ORPHE ANALYTICS has not yet been used to verify how consistent the position/angle-level gait parameters calculated using this motion-sensor-based system and its software are with the corresponding parameters obtained using the conventional optical motion capture.

Therefore, this study aimed to validate the reliability of ORPHE ANALYTICS by assessing the agreement of position/angle-level gait parameters between ORPHE ANALYTICS and optical motion capture during walking and running in healthy participants. We evaluated the relative validity between the gait parameters from the two different modalities using intraclass correlation coefficients (ICCs) and visualised them using Bland–Altman plots. We hypothesised that the gait parameters calculated using ORPHE ANALYTICS exhibit excellent relative validities (ICC>0.9) to those calculated using optical motion capture.

## Methods

### Participants

Nine healthy volunteers who had not undergone surgical treatment of the lower extremity within one year before the experiment participated in the study [six men and three women; mean age: 25.4 (2.2) y; mean height: 166.6 (9.7) cm; mean weight: 60.3 (10.7) kg; mean shoe size: 25.7 (1.3) cm]. A priori power analysis suggested a required trial sample size of N=117, with power=0.8, alpha=0.05, null assumption of ICC=0.7, and alternative hypothesis=0.8, based on the formula provided by Zou[13]. The Osaka University Hospital Ethics Committee approved this study (approval no. 19537), and all participants provided informed consent.

### Procedure

The participants wore shoes (SHIBUYA 2.0, ORPHE Inc., Tokyo, Japan) with midsole space to embed the motion sensor (ORPHE Inc., Tokyo, Japan; size: 45 mm×29 mm×14 mm, weight: 20 g; Figure 1a). This motion sensor samples the three-axial accelerations and angular velocities using a built-in sensor (LSM6DSOX, STMicroelectronics; acceleration: ± 16 G, angular velocity: ± 2000 °/s) at a sampling frequency of 200 Hz. The recorded data were stored in built-in flash memory. Two motion sensors were utilised for each foot. One sensor was embedded in the midsole space (plantar-embedded, Figure 1a), and the other was securely fixed onto the shoe instep using dedicated mounting equipment (instep-mounted, Figure 1b). Fourteen reflective markers were attached to the shoe landmarks to specify the posture and position of the motion sensor on the foot (Figure 1c).

**Figure 1.**
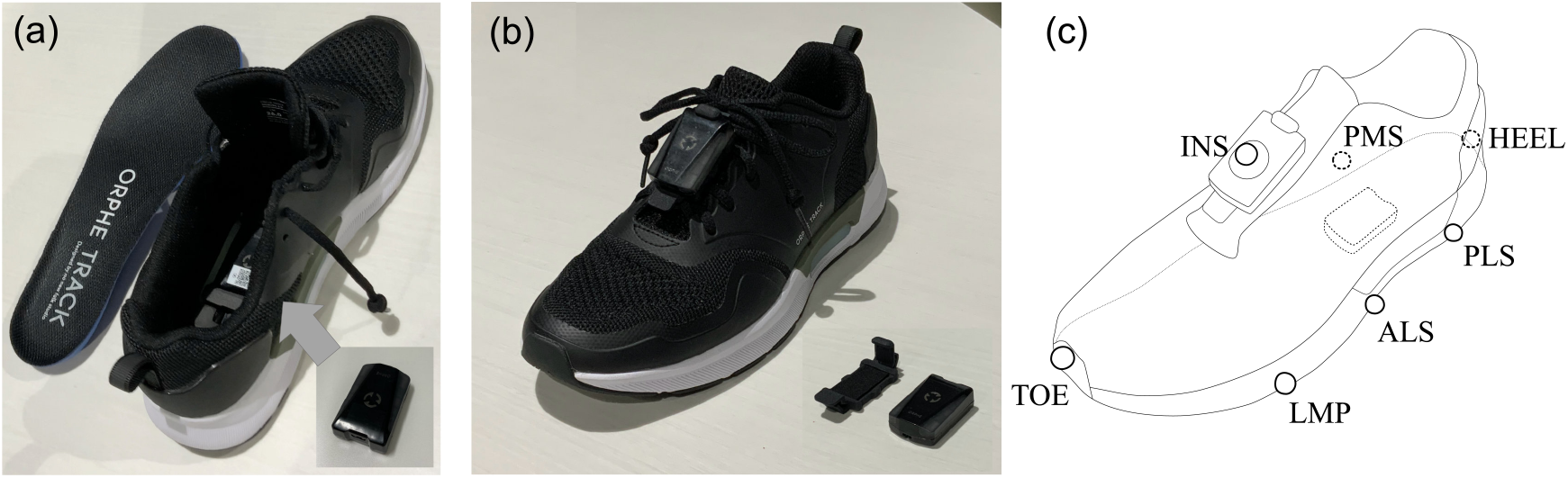
(a) Motion sensor embedded in the shoe midsole; (b) Motion sensor mounted on the shoe instep and dedicated mounting equipment; (c) Marker position on the shoe (a) The shoe has space for sensor placement, and the centre of the sensor locates 40% of the shoe length from the heel edge. (b) Motion sensor mounted on the shoe instep using dedicated mounting equipment that can be fixed to the shoelace. (c) The names and position definitions of the reflective markers are as follows: LMP, the lateral edge of the metatarsophalangeal joint; TOE, most anterior edge of the shoe; PLS, posterior lateral side of motion sensor; ALS, anterior lateral side of motion sensor; PMS, posterior medial side of motion sensor; HEEL, most posterior edge of shoe midsole; INS, on the centre of the sensor mounted on instep equipment of the shoe.

Participants performed walking and running tasks on a treadmill (MyRun, Technogym, Cesena, Italy). The walking task required double support phases and was performed at target speeds of 2, 4, and 6 km/h. The running task required no double support phases and was performed at target speeds of 6, 9, and 12 km/h. The three foot-strike conditions, that is, the self-selected foot, forefoot, and rearfoot strike patterns, were performed only in the 12-km/h running task. The different foot-strike patterns’ purpose was to replicate natural running variability and increase the associated data variation. Foot-strike pattern differences have been reported to affect gait-event detection timing in gait analysis using foot-mounted motion sensors[14]. Two experimenters (UY and YN) monitored task requirements. At the beginning of each trial, the participant maintained a static standing position on the treadmill for 10 s and subsequently performed a vertical jump to synchronise the motion sensors and motion capture system. Thereafter, the participants were asked to increase the treadmill speed by themselves to the target speed and maintain the assigned gait speed for 1 min. Tasks were performed from the slower to the faster condition for the participants’ safety. Three-minute resting periods were provided between each trial to reduce the effect of fatigue. The participants could discontinue their trial at any time when they experienced difficulty in completing further trials (see Supplementary Table S1 for details of the tasks completed by the participants). The reflective-marker positions were measured using 12 optical cameras (OptiTrack Prime 17 W, NaturalPoint, Inc., Corvallis, OR, USA) at a 360-Hz sampling frequency. The three-axial accelerations and angular velocities of the four motion sensors (plantar/instep on both sides) were sampled at 200 Hz.

### Data processing

The marker position data were smoothed using a second-order Butterworth digital filter (low-pass, zero-lag, cut-off frequency:10 Hz). The motion sensor centre position, anteroposterior axis, and mediolateral axis used in parameter calculation were defined as follows. The motion sensor centre was the midpoint between the ALS and PMS markers. The anteroposterior axis was defined as the unit vector extending from PLS to the ALS marker. The support vector was defined as the unit vector extending from the PMS to the PLS for the right foot and from the PLS to the PMS for the left foot. The vertical axis was defined as the cross-product of the support vector and anteroposterior axis vector. The mediolateral axis was defined as the cross-product of the anteroposterior and vertical axis vectors.

The timing of the initial foot contact (IC) for each gait cycle was defined as the instance at which the target marker’s peak vertical acceleration appeared[15]. The target marker was selected in every step according to the foot’s orientation when the vertical distance between any of the foot markers and the treadmill surface was < 50 mm. The HEEL marker was assigned as the target marker when the angle between the motion sensor’s anteroposterior axis and global horizontal plane was < −15° (toe-up), the LMP marker was assigned when the angle ranged from −15° to −5° (near-flat), and TOE marker was assigned when the angle was > −5° (toe-down) (Figure S1). The timing of the toe-off (TO) was defined when the TOE marker exceeded 10 mm above the minimum vertical height for each cycle[16]. One gait cycle was defined from one IC to the next for each foot[17]. The gait parameters listed in Table 1 were calculated using each gait cycle’s motion capture data.

**Table 1.** Gait parameters and their definitions

| Parameter name | Unit | Explanation | Definition of motion capture data analysis | Name in ORPHE ANALYTICS* |
| --- | --- | --- | --- | --- |
| Stride duration | sec | A time required for 1 gait cycle | A time required for IC to next IC | Stride duration |
| Stride length | m | Anteroposterior displacement of the foot during 1 gait cycle | Difference between the anteroposterior position of the motion sensor centre at IC and next IC + (stride duration) * (treadmill speed) | Stride length |
| Stride frequency | steps/sec | Number of gait cycles per 1 second | The inverse of stride duration | Stride frequency |
| Stride speed | m/sec | Average speed of foot during 1 gait cycle | Stride length / stride duration | Stride speed |
| Vertical height | m | Maximum height of foot during 1 gait cycle | Maximum height of the centre of motion sensor during 1 gait cycle | Vertical height |
| Stance phase duration | sec | A time that the foot is in contact with the ground during 1 gait cycle | Time required for IC to next TO | Ground contact time |
| Swing phase duration | sec | A time that the foot is in the air during 1 gait cycle | Time required for TO to next IC | Swing time |
| Sagittal angle<sub>IC</sub> | degree | The angle between ground and foot in the sagittal plane at IC | The angle between the motion sensor anteroposterior axis and its projection vector to the horizontal plane at IC | Strike angle |
| Frontal angle<sub>IC</sub> | degree | The angle between ground and foot in the frontal plane at IC | The angle between the motion sensor mediolateral axis and its projection vector to the horizontal plane at IC | Pronation |
| Sagittal angle<sub>TO</sub> | degree | The angle between ground and foot in the sagittal plane at TO | The angle between the motion sensor anteroposterior axis and its projection vector to the horizontal plane at TO | Toe off angle |

Note: IC: initial foot contact; TO: toe-off; \*: cited at https://orphe.io/en/analytics.

The gait parameters based on the motion sensor data were calculated using ORPHE ANALYTICS (version 1.4.2; ORPHE Inc., Tokyo, Japan) for each gait cycle (Table 1).

We defined the gait cycles to be analysed and calculated the gait-cycle detection ratio because ORPHE ANALYTICS did not detect the gait cycle ideally. The motion sensor and optical motion capture data were time-synchronised with the initial vertical jump’s landing timing, as described in the method. This procedure was not exact-time synchronisation but sufficient for gait-cycle-wise correspondence between modalities. To analyse the period after the treadmill speed reached the target speed, the first 15 gait cycles detected from the motion sensor data were discarded. The period from 350 ms before the 16th gait cycle to 350 ms after the last gait cycle was defined as the analysis period. Regarding the gait cycles calculated from the optical motion capture data, the gait cycles covered in the analysis period were analysed. The gait-cycle detection ratio was calculated as the number of gait cycles for analysis by the motion sensor divided by that for analysis by the optical motion capture.

Outlier processing was performed to exclude gait cycles containing abnormal gait-parameter values after calculating the gait-cycle detection ratio. Quartiles were calculated for each trial based on the stride length and duration obtained from the motion sensor and motion capture data. Gait cycles that included stride length or duration outside 1.5 times the interquartile range above the upper quartile point or below the lower quartile point were excluded from the analysis. The percentage of excluded gait cycles was subsequently calculated.

### Statistical analysis

The average gait-parameter values for each trial were calculated separately for the left and right sides, and statistical processing was performed. To evaluate the relative validity of the gait parameters calculated using motion sensor data against that of those calculated using the motion capture, ICC_(2,1)_ values were calculated for the overall (walking and running), walking, and running conditions. ICC_(2,1)_ values were interpreted as follows: excellent (>0.90), good (0.75-0.90), moderate (0.50-0.75), and poor (<0.50)[18].

To visualise the bias and precision between the gait parameters from the motion sensor data and those from the motion capture data, Bland–Altman plots[19] were plotted. Statistical indices, such as mean difference, 95% limits of agreement of the difference (LoA_95%_), and percentage LoA (LoA%), in the Bland–Altman plot were also calculated for the overall, walking, and running conditions to evaluate absolute reliability. The LoA% was interpreted as follows: very good (<5%), good (5–10%), moderate (10–20%), poor (20–50%), and very poor (>50%)[20]. However, the LoA% of the sagittal angle_IC_ and frontal angle_IC_ could not be calculated because these gait parameters’ average values were normally distributed around 0, rendering it inappropriate to evaluate in percentages.

## Results

### Gait-cycle detection ratio in ORPHE ANALYTICS

The status of trial execution and data acquisition is shown in Supplementary Table S1. As regards the plantar-embedded motion sensor, the gait-cycle frequencies detected by the motion sensor and optical motion capture data were 5,931 and 5,981, respectively, and the gait-cycle detection ratio was 99.16%. Regarding the instep-mounted motion sensor, the gait-cycle frequencies detected by the motion sensor and optical motion capture data were 6,343 and 6,372, respectively, and the gait-cycle detection ratio was 99.54% (Supplementary Table S2).

### Gait-cycle percentages including outliers

The percentages of gait-cycle data excluded from the analysis as outliers were 10.84% and 9.52% for the plantar-embedded and instep-mounted motion sensors, respectively (Supplementary Table S3).

### Relative validity of ORPHE ANALYTICS against optical motion capture

The gait-parameter ICCs from ORPHE ANALYTICS and optical motion capture are shown in Table 2. Stride length, stride duration, stride frequency, stride speed, vertical height, stance phase duration, swing phase duration and sagittal angle_IC_ exhibited excellent relative validities (ICC>0.9) in both plantar-embedded and instep-mounted motion sensors. The frontal angle_IC_ demonstrated moderate agreement (ICC=0.566–0.627), while the sagittal angle_TO_ exhibited good agreement (ICC=0.892–0.833).

**Table 2.**
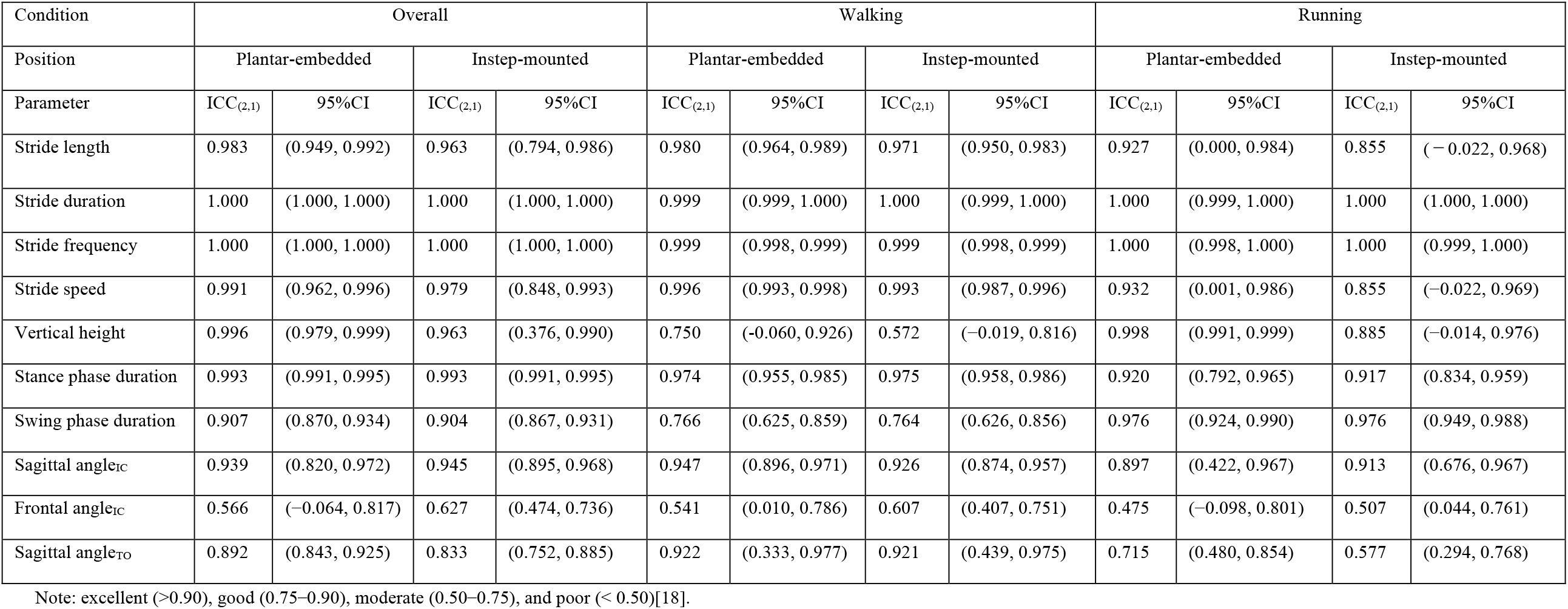
Intraclass correlation coefficients of the gait parameters from ORPHE ANALYTICS and optical motion capture

### Absolute reliability of ORPHE ANALYTICS against optical motion capture

The Bland–Altman plots of the gait parameters from ORPHE ANALYTICS and the optical motion capture are shown in Figures 2 and 3, and the statistical indices are shown in Table 3. Stride duration, stride frequency, vertical height (plantar-embedded), and stride speed (plantar-embedded) exhibited very good-to-good absolute reliability (LoA%=1.0–9.8). Stride length, stride speed (instep-mounted), vertical height (instep-mounted), stance phase duration, swing phase duration, and sagittal angle _TO_ demonstrated moderate-to-poor absolute reliability (LoA%=10.0–20.4). The vertical height’s LoA% was extremely poor for the instep-mounted model compared with that for the plantar-embedded model (Table 3). Proportional errors were visually recognised for stride length and speed (Figure 2b,d, Figure 3b,d).

**Table 3.** Statistical indices of the Bland–Altman plots

| Condition | Overall |  |  |  | Walking |  |  |  | Running |  |  |  |
| --- | --- | --- | --- | --- | --- | --- | --- | --- | --- | --- | --- | --- |
| Position | Plantar-embedded |  | Instep-mounted |  | Plantar-embedded |  | Instep-mounted |  | Plantar-embedded |  | Instep-mounted |  |
| Parameter | Mean diff. ±<br>LoA95% | LoA% | Mean diff. ±<br>LoA95% | LoA% | Mean diff. ±<br>LoA95% | LoA% | Mean diff. ±<br>LoA95% | LoA% | Mean diff. ±<br>LoA95% | LoA% | Mean diff. ±<br>LoA95% | LoA% |
| Stride length (m) | $-0.058 \pm 0.174$ | 10.0 | $-0.105 \pm 0.224$ | 13.1 | $0.016 \pm 0.103$ | 8.1 | $-0.003 \pm 0.127$ | 10.1 | $-0.111 \pm 0.135$ | 6.5 | $-0.176 \pm 0.165$ | 8.1 |
| Stride duration (s) | $-0.002 \pm 0.013$ | 1.3 | $-0.001 \pm 0.011$ | 1.1 | $-0.003 \pm 0.019$ | 1.5 | $-0.003 \pm 0.016$ | 1.3 | $-0.001 \pm 0.003$ | 0.4 | $-0.001 \pm 0.004$ | 0.5 |
| Stride frequency (steps/s) | $0.001 \pm 0.012$ | 1.1 | $0.001 \pm 0.012$ | 1.0 | $-0.000 \pm 0.017$ | 2.0 | $0.000 \pm 0.017$ | 2.0 | $0.002 \pm 0.006$ | 0.4 | $0.001 \pm 0.006$ | 0.4 |
| Stride speed (m/s) | $-0.081 \pm 0.200$ | 9.8 | $-0.14 \pm 0.275$ | 13.6 | $0.006 \pm 0.075$ | 6.9 | $-0.012 \pm 0.098$ | 9.0 | $-0.142 \pm 0.172$ | 6.4 | $-0.228 \pm 0.219$ | 8.2 |
| Vertical height (m) | $-0.009 \pm 0.020$ | 6.6 | $0.027 \pm 0.035$ | 20.4 | $-0.016 \pm 0.018$ | 10.9 | $0.009 \pm 0.015$ | 20.4 | $-0.004 \pm 0.013$ | 3.4 | $0.040 \pm 0.020$ | 8.4 |
| Stance phase duration (m) | $-0.003 \pm 0.069$ | 13.1 | $-0.005 \pm 0.069$ | 13.3 | $-0.009 \pm 0.097$ | 11.5 | $-0.011 \pm 0.095$ | 11.3 | $0.002 \pm 0.036$ | 11.9 | $-0.001 \pm 0.04$ | 13.4 |
| Swing phase duration | $0.002 \pm 0.065$ | 14.5 | $0.004 \pm 0.065$ | 14.5 | $0.007 \pm 0.090$ | 21.9 | $0.010 \pm 0.088$ | 21.5 | $-0.002 \pm 0.036$ | 7.6 | $0.000 \pm 0.040$ | 8.4 |
| Sagittal angle<sub>IC</sub> (°) | $-2.245 \pm 6.355$ | N/A | $-1.609 \pm 6.459$ | N/A | $-0.937 \pm 4.428$ | N/A | $-0.752 \pm 5.100$ | N/A | $-3.171 \pm 6.868$ | N/A | $-2.203 \pm 7.021$ | N/A |
| Frontal angle<sub>IC</sub> (°) | $3.849 \pm 5.756$ | N/A | $1.551 \pm 7.770$ | N/A | $2.858 \pm 5.528$ | N/A | $-0.784 \pm 5.560$ | N/A | $4.551 \pm 5.513$ | N/A | $3.168 \pm 7.498$ | N/A |
| Sagittal angle<sub>TO</sub> (°) | $-1.107 \pm 7.868$ | -12.6 | $-1.701 \pm 9.691$ | -15.6 | $-3.124 \pm 4.867$ | -7.3 | $-2.994 \pm 5.199$ | -7.8 | $0.321 \pm 8.371$ | -14.2 | $-0.806 \pm 11.519$ | -19.4 |

Note: Mean diff.: mean difference, N/A: not available (the ICCs of sagittal angle<sub>IC</sub> and frontal angle<sub>IC</sub> were not calculated because those values were distributed positively and negatively, and those averages were close to 0, making it inappropriate to evaluate in percentages.). The LoA% was interpreted as follows: very good (<5%), good (5–10%), moderate (10–20%), poor (20–50%), and very poor (>50%)[20].

**Figure 2.**
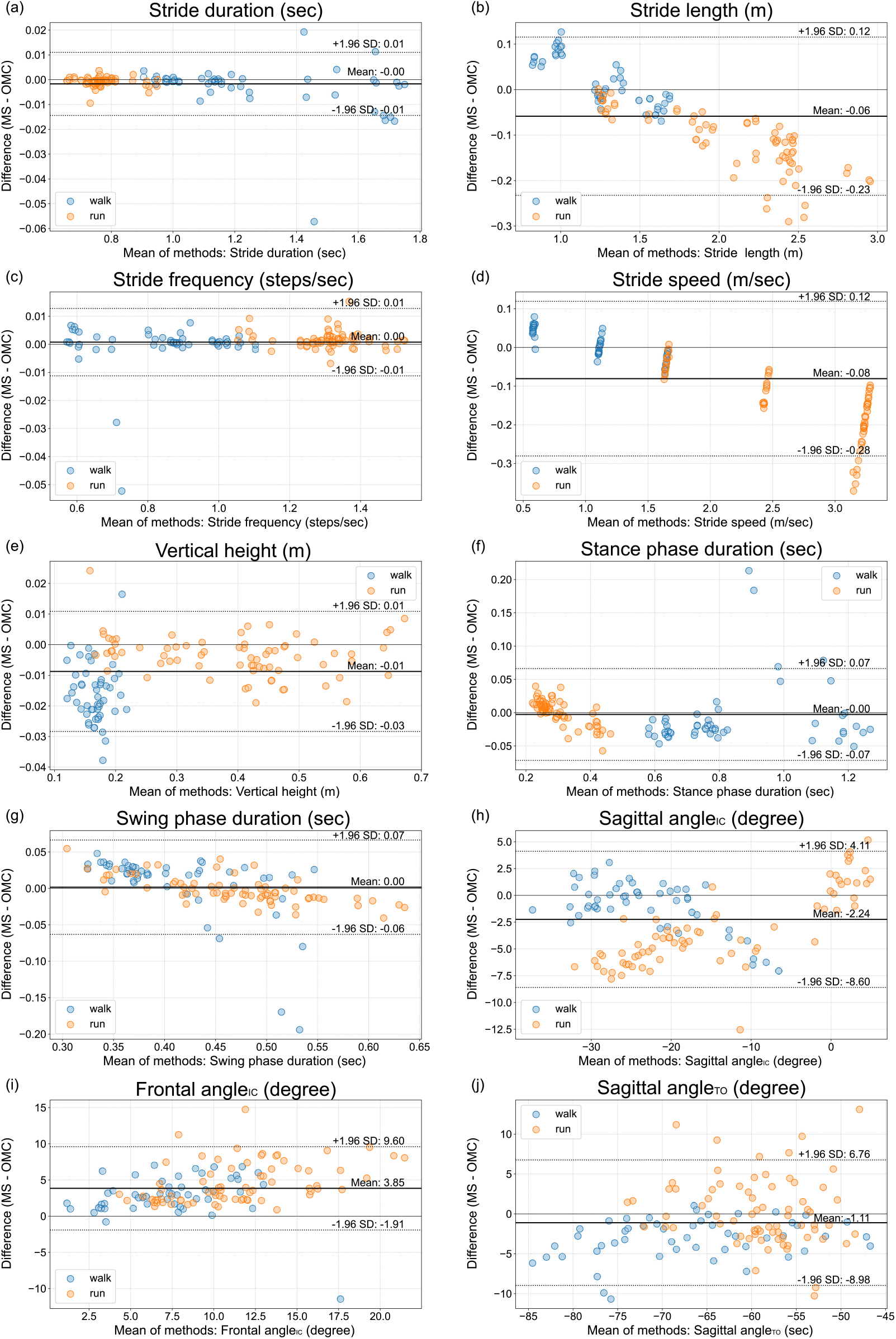
Bland–Altman plot of gait parameters from the motion sensor embedded in the shoe midsole (plantar-embedded) and optical motion capture (a) stride duration, (b) stride length, (c) stride frequency, (d) stride speed, (e) vertical height, (f) stance phase duration, (g) swing phase duration, (h) sagittal angle_IC_, (i) frontal angle_IC_, (j) sagittal angle_TO_ MS: motion sensor, OMC: optical motion capture

**Figure 3.**
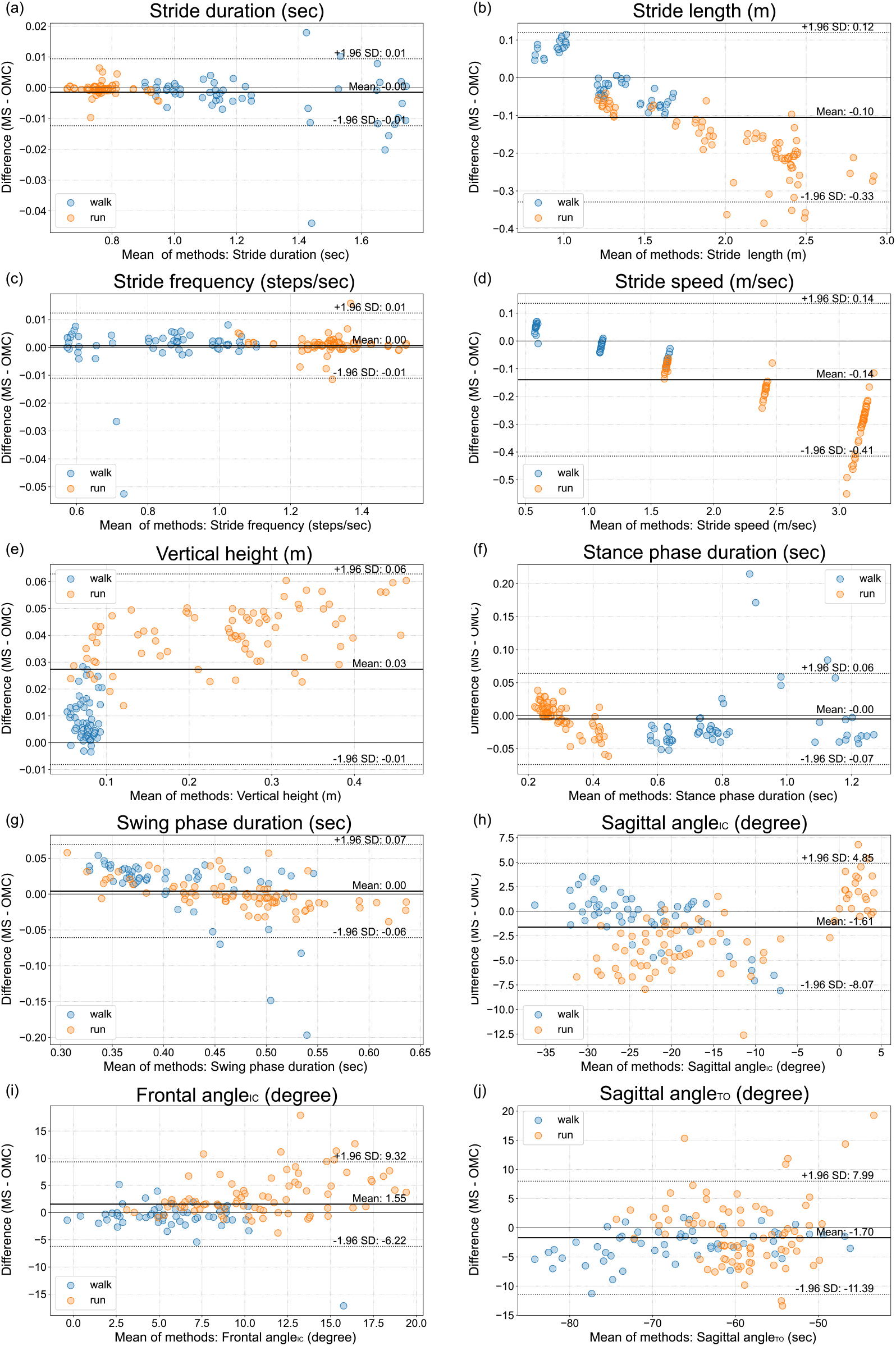
Bland–Altman plot of gait parameters from the motion sensor mounted on the shoe instep (instep-mounted) and optical motion capture (a) stride duration, (b) stride length, (c) stride frequency, (d) stride speed, (e) vertical height, (f) stance phase duration, (g) swing phase duration, (h) sagittal angle_IC_, (i) frontal angle_IC_, (j) sagittal angle_TO_ MS: motion sensor, OMC: optical motion capture

## Discussion

In this study, ORPHE ANALYTICS-derived gait parameters were compared with those from optical motion capture data during walking and running tasks at speeds of 2–12 km/h in nine healthy adults. The relative validities of the stride length, stride duration, stride frequency, stride speed, vertical height, stance phase duration, swing phase duration, and sagittal angle_IC_ for both plantar-embedded and instep-mounted motion sensors were excellent (ICC>0.9) with respect to the optical motion capture (Table 2); however, those of the sagittal angle_TO_ and sagittal angle_IC_ were good, while those of the frontal angle_IC_ were moderate. These results partially support our hypothesis.

The relative validities of ORPHE ANALYTICS-derived gait parameters were not low compared with those of a previous meta-analysis involving commercial products such as Physilog, Mobility Lab, and Stryd, among others (ICC: stance phase time, 0.81–0.97; swing phase time, 0.56–0.81; stride duration, 0.55–0.99; stride frequency, 0.96–0.99; stride length, 0.75–0.99)[21]. The Bland– Altman plots revealed that stride length and speed had proportional relationships, and as the values increased, those calculated using ORPHE ANALYTICS became smaller than those calculated using the optical motion capture. This study’s results, based on treadmill use by healthy adults, endorse the reliability of ORPHE ANALYTICS in clinical applications.

### Differences between plantar-embedded and instep-mounted motion sensors

Regarding the mounting position, ORPHE ANALYTICS’ validity relative to optical motion capture was generally better for the plantar-embedded than for the instep-mounted motion sensor (Tables 2 and 4). Major differences were observed primarily in vertical height, stride length, stride speed, and sagittal angle_TO_. A previous study reported that the error against the reference was smaller for a sensor embedded in the shoe midsole than for that mounted on the shoe instep[12,22]. This was due to an error in the integral calculation caused by the relatively high vibration susceptibility of the sensor mounted on the shoe instep[12]. Although we used dedicated mounting equipment to fix the motion sensor to the shoelace, complete suppression of the influence of vibration was difficult. While dedicated mounting equipment is advantageous in that the sensor can be attached to a variety of shoe types, the original shoes with space for storing the sensor have the advantage of higher gait analysis validity.

### Less absolute reliability in sagittal angle_TO_, swing phase duration, and stance phase duration

The Bland–Altman plots revealed relatively large LoA% in swing phase duration during walking as well as stance phase duration and sagittal angle_TO_ during running (Table 3). These errors were caused by differences in toe-off timing detection. Toe-off timing detection accuracy directly influences these parameters. Due to walking’s short swing phase duration, the time deviation of the toe-off timing detection relative to the swing phase duration becomes larger. Hence, the LoA% of the swing phase duration during walking is more prominent, exhibiting a trend similar to that in previous studies[6,23]. Conversely, the LoA% of the swing phase duration was slightly smaller during running because the swing phase duration was relatively longer. Even in previous motion-sensor-based gait analyses, the rule-based detection of toe-off timing from motion sensor data has been reported to be complex and inaccurate[14,24]. The difficulty in detecting toe-off timing may also influence the large absolute error of the sagittal angle_TO_ during running.

### The proportional errors in stride speed and stride length

As the treadmill speed increased, the ORPHE ANALYTICS-derived stride speed and length tended to be smaller than those calculated using optical motion capture data (Figures 2b, 2d, 3b, 3d). These results are consistent with those reported in previous studies that analysed running using motion sensors[12,25]. Falbriard et al. [25] noted certain limitations in estimating stride speed through the simple integration of accelerations during running. They attempted a correction using a linear function for the stride speed estimated by integrating 500-Hz sampling frequency accelerations. Considering these arguments, we suggest the following caution when interpreting the values. First, high relative validity implies its suitability for explaining changes in relative values, for example, evaluating changes in the same-person’s speed. However, to refer to absolute values, a better estimation can be made by correcting for proportional agreement. When stride speed and length were corrected based on the regression lines obtained from this study’s results, the LoA% decreased and exhibited excellent reliability (Supplementary Figure S2). Interpreting stride speed and length using ORPHE ANALYTICS based on the assumption of existent proportional differences is necessary.

### Limitations

This study had certain limitations. ORPHE ANALYTICS validation was conducted using a treadmill because a conventional optical motion capture was used for comparison, and obtaining many samples for walking and running was necessary. In many clinical situations, treadmill-based gait is used as a task and measured using motion capture to measure speed-controlled data. In our validation, we initially provided a standard for the comparison of ORPHE ANALYTICS with optical motion capture data during treadmill-based walking and running.

Only healthy young adults participated in this study. To examine a wide range of gait speeds and foot-strike patterns, we selected healthy young adult participants with a high potential for exercise. However, gait patterns vary with age and disease status. Further validation is necessary for older people and patients with diseases whose gait patterns may affect gait-parameter calculation using motion sensors[6].

## Conclusion

In this study, we evaluated the relative validity and absolute reliability of gait parameters calculated using the motion-sensor-based gait analysis system ORPHE ANALYTICS against those calculated using optical motion capture in 2–12 km/h gait involving nine healthy young adults.

Stride duration, stride length, stride frequency, stride speed (plantar-embedded), vertical height (plantar-embedded), stance phase duration, swing phase duration, and sagittal angle_IC_ exhibited excellent relative validity. However, sagittal angle_TO_ and frontal angle_IC_ exhibited good and moderate relative validities, respectively.

ORPHE ANALYTICS enables gait analysis regardless of the measurement environment and is expected to be applied in daily-life measurements. This study’s results will serve as a reliability standard for the future use of this gait analysis system.

## Data Availability

All data produced in the present study are available upon reasonable request to the authors

## Acknowledgement

This work was supported by MEXT “Innovation Platform for Society 5.0” Program Grant Number JPMXP0518071489. The funder had no role in the study design, collection, analysis, and interpretation of data, manuscript preparation, and decision to submit the article for publication.

## Supplementary Material

**Supplementary Table S1.** Status of trial execution and data acquisition

| ID | sex | Walking |  |  | Running |  |  |  |  |
| --- | --- | --- | --- | --- | --- | --- | --- | --- | --- |
|  |  | 2km/h | 4km/h | 6km/h | 6km/h | 9km/h | 12km/h | 12km/h forefoot strike | 12km/h rearfoot strike |
| 1 | Male | ● | ● | ● | ● | ● | ● | ● | ● |
| 2 | Female | ● | ● | ● | ● | ● |  |  |  |
| 3 | Female | † | ● | ● | ● | ● | ● |  |  |
| 4 | Female | ● | ● | ● | ● | ● | ● | ● | ● |
| 5 | Male | ● | ● | ● | ● | ● | ● | ● | ● |
| 6 | Male | ● | ● | ● | ● | ● | ● | ● | ● |
| 7 | Male | ● | ● | ● | ● | ● | ● | ● | ● |
| 8 | Male | ● | ● | ● | ● | ● | ● | † | ● |
| 9 | Male | ● | * | * | * | * | * | * | * |

●: Data with no deficits; †: Deficient left plantar-embedded sensor data. \*: Deficient right instep-mounted sensor data; no marker: the trial was not executed.

**Supplementary Table S2.**
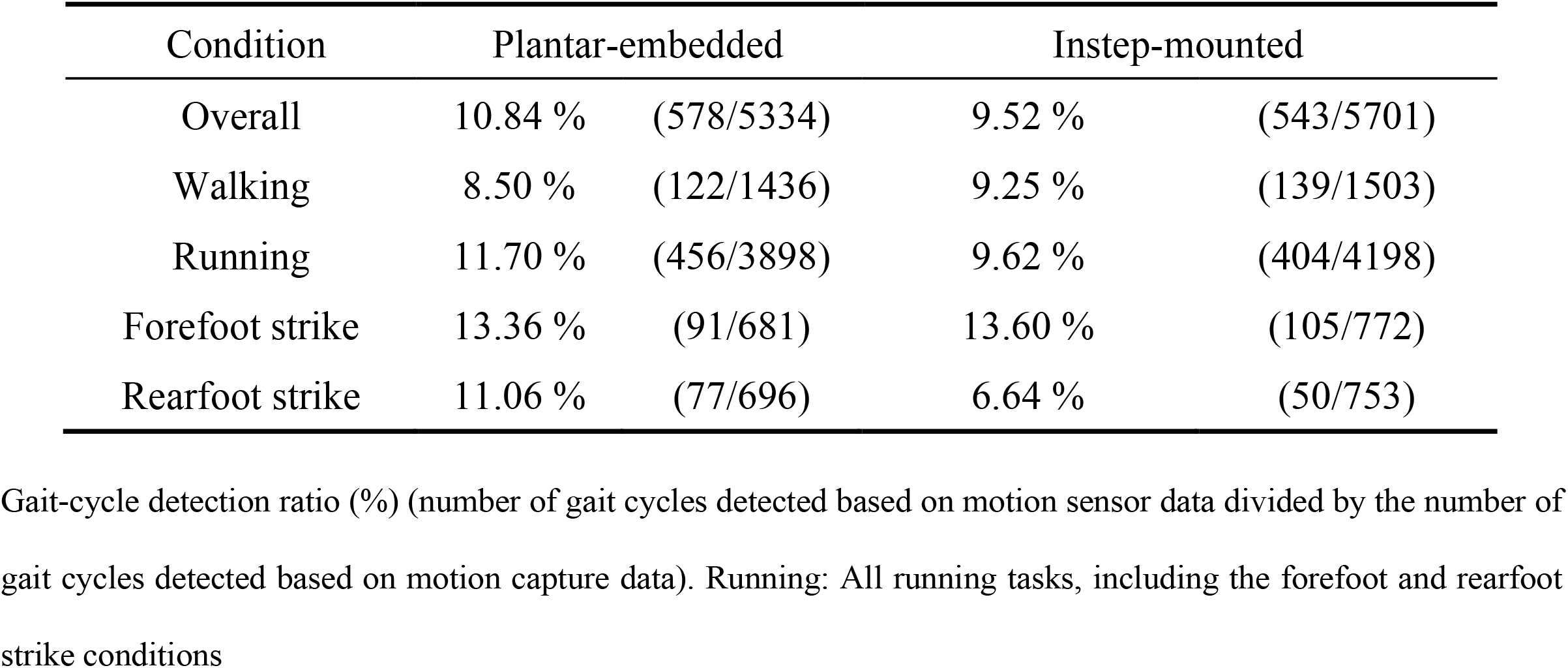
Gait-cycle detection ratio for the conditions

**Supplementary Table S3.**
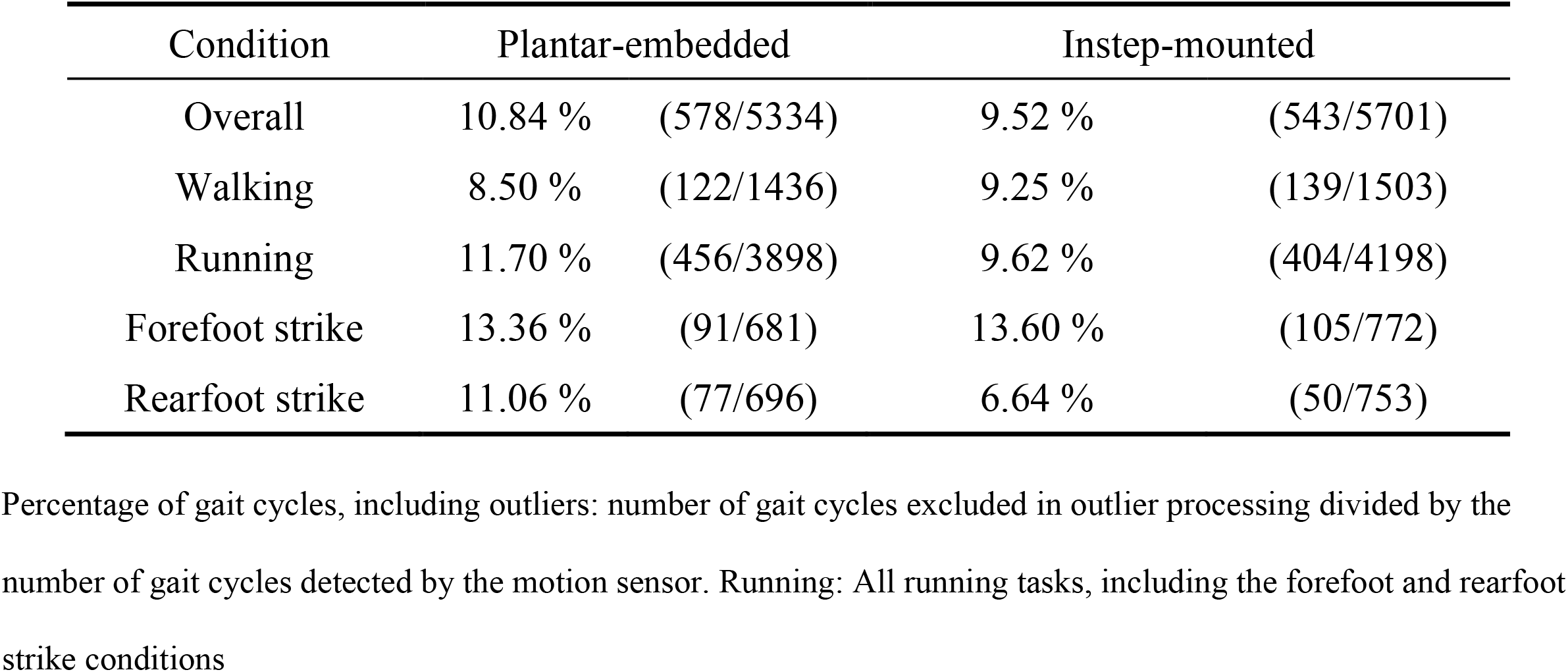
Percentage of gait cycles, including outliers for each condition

**Supplementary Figure S1.**
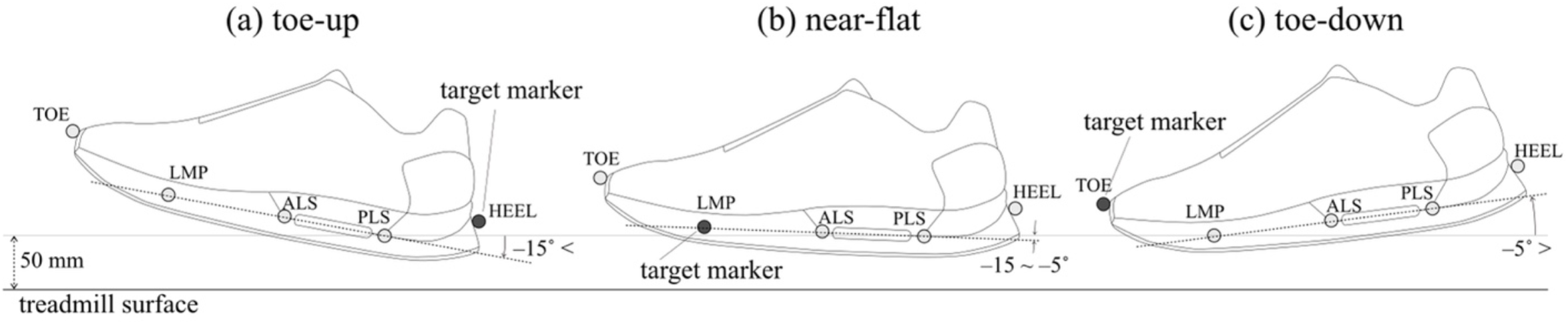
Definition of target markers for detecting the initial foot contact timing. (a) The HEEL marker was assigned as the target marker when the angle between the anteroposterior axis of the motion sensor and global horizontal plane was less than −15° (toe-up). (b) The LMP marker was assigned when the angle ranged from −15° to −5° (near-flat). (c) The TOE marker was assigned when the angle was more than −5° (toe-down).

**Supplementary Figure S2.**
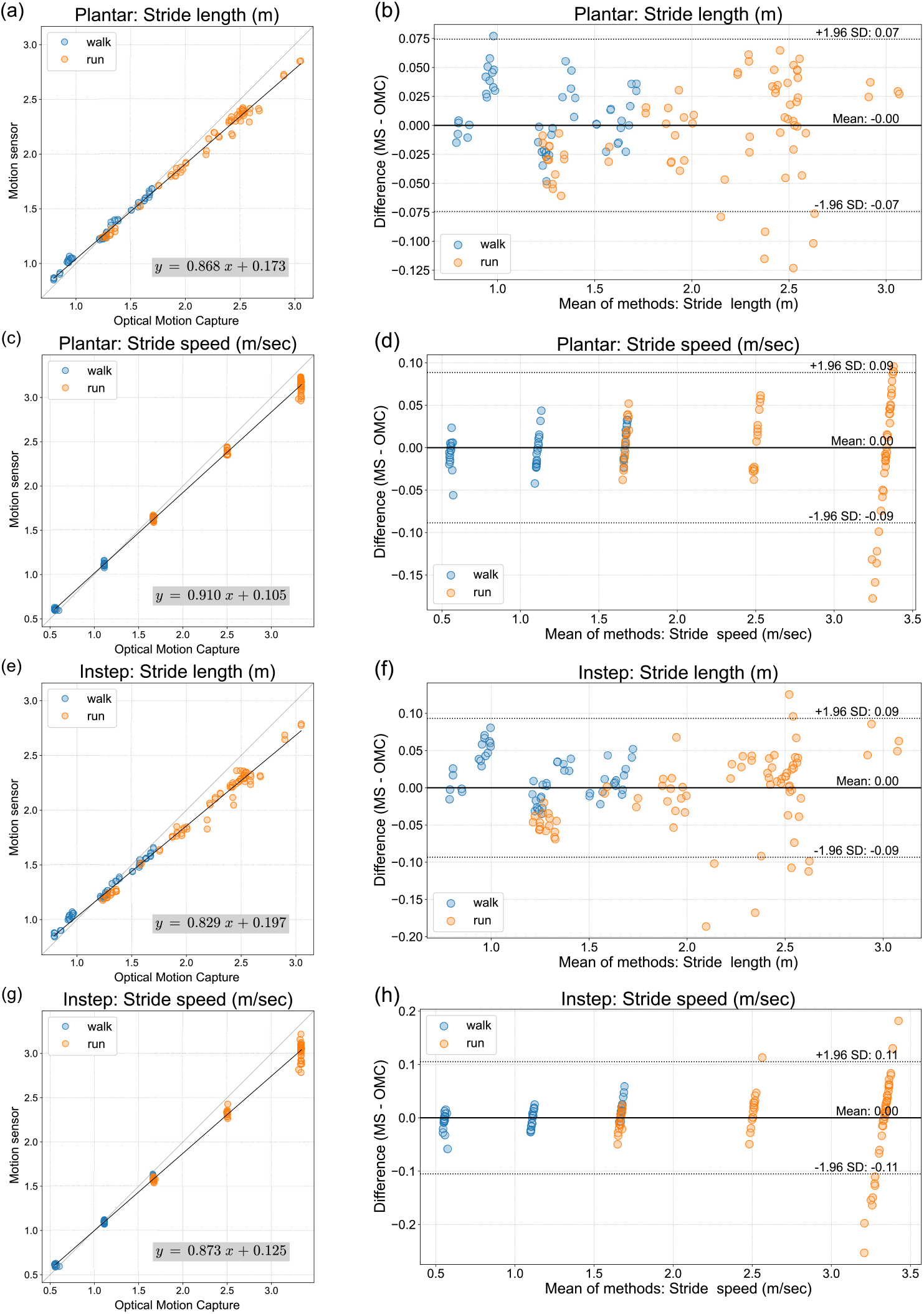
Scatter plots and regression lines of the stride length and speed from ORPHE ANALYTICS and the optical motion capture as well as the Bland–Altman plots of corrected data using regression lines. (a) Scatter plot: stride length (plantar); (b) Bland–Altman plot: stride length (plantar); (c) scatter plot: stride speed (plantar); (d) Bland–Altman plot: stride speed (plantar); (e) scatter plot: stride length (instep); (f) Bland–Altman plot: stride length (instep); (g) scatter plot: stride length (instep); (h) Bland–Altman plot: stride length (instep). MS: motion sensor, OMC: optical motion capture

